# ClearSpeechTogether: a rater blinded, single, controlled feasibility study of speech intervention for people with progressive ataxia

**DOI:** 10.1101/2022.04.19.22273510

**Authors:** Anja Lowit, Jessica Cox, Melissa Loucas, Jennifer Grassly, Aisling Egan, Frits van Brenk, Marios Hadjivassiliou

## Abstract

**Background:** Progressive ataxias frequently lead to speech disorders and consequently impact on communication participation and psychosocial wellbeing. Whilst recent studies demonstrate the potential for improvements in these areas, these treatments generally require intensive input which can reduce acceptability of the approach.

A new model of care – ClearSpeechTogether – is proposed which maximises treatment intensity whilst minimising demands on clinician. This study aimed to establish feasibility and accessibility of this approach and at the same time determine the potential benefits and adverse effects on people with progressive ataxias.

**Method:** The study targeted people with progressive ataxia and mild-moderate speech and gross motor impairment. ClearSpeechTogether consisted of four individual sessions over two weeks followed by 20 patient-led group sessions over four weeks. All sessions were provided online. Quantitative and qualitative data were collected for evaluation.

**Results:** Nine participants completed treatment. Feasibility and acceptability were high and no adverse effects were reported. Statistical tests found significantly reduced vocal strain, improved intelligibility for reading, and increased participation and confidence. Participant interviews highlighted the value of group support, from psychosocial perspectives and in supporting speech strategy internalisation and generalisation.

**Discussion:** ClearSpeechTogether presented an effective intervention in a small group of people with progressive ataxia. It matched or exceeded the outcomes previously reported for intensive, individual therapy while minimising clinician time demands. Furthermore, its unique peer led group intervention design appeared effective in addressing intractable psychosocial issues. ClearSpeechTogether is potentially cost-effective, providing intensive delivery with few clinician sessions, thus maximising the input available from health care providers.

## Introduction

Progressive ataxia is the term used to describe a number of different diseases that primarily affect the cerebellum resulting in loss of co-ordination, limb clumsiness, gait instability, falls, slurred speech and sometimes visual problems. The term tends to be used in the context of those ataxias that are not due to a structural pathology (e.g., tumour, stroke, multiple sclerosis, trauma). As the majority of ataxias are not treatable, patients accumulate significant disability over time, sometimes becoming wheel-chair dependant with reduced lifespan. The causes of progressive ataxias can be broadly divided into genetic, acquired (non-degenerative) and degenerative. The commonest inherited ataxia is Friedreich’s ataxia (FRDA). The prevalence of other genetic ataxias has considerable geographical variation. Non-degenerative acquired ataxias include immune ataxias (e.g., gluten ataxia, paraneoplastic cerebellar degeneration, post infectious cerebellitis), and the most notable example of a degenerative ataxia is cerebellar variant of multiple system atrophy (MSA-C). Depending on the aetiology, ataxias can progress rapidly (e.g., immune ataxias and MSA-C) or slowly over many years (e.g., genetic ataxias). Depending on aetiology some ataxias can be more commonly associated with dysarthria (e.g., MSA-C) whilst others can be more commonly associated with gait instability (e.g., immune ataxias).

As the disease progresses ataxia can lead to speech problems, presenting as ataxic dysarthria. The nature and onset of disease varies across and even within ataxia types. For example, Friedreich’s ataxia (FRDA) has been described as falling into three different categories of progression and symptomatology [1], and the same is true for dominantly inherited spino-cerebellar ataxias (SCA) [2, 3]. Despite these individual differences, ataxic dysarthria can generally be characterised by symptoms impacting on all speech sub-systems, i.e., respiration, laryngeal function, articulation and resonance [4-16], leading to reduced speech intelligibility and communication breakdown. This is likely to have further consequences on an individual’s quality of life. Studies on communication impact of dysarthria due to other neurological conditions such as Parkinson’s Disease or following stroke suggest that speakers experience poor mental health, negative self-image and withdrawal from communication and thereby social contacts [17-20]. No such studies have been published for people with ataxia, however, more than a third of respondents to a recent survey by Ataxia UK identified speech problems as one of the three most troublesome symptoms of their disease [21]. Whilst our understanding of the nature of the communication problems experienced by people with progressive ataxia has improved significantly over time [1, 5, 7, 9, 12-15], a 2017 Cochrane Review on treatment efficacy for these syndromes concluded that insufficient and low quality evidence was available on the effectiveness of speech interventions to support this population [22].

Since the publication of the Cochrane review, further studies relating to speech treatment in progressive ataxia syndromes have been published. Together these studies have highlighted a range of potential communication benefits across all areas of the International Classification of Functioning and Disability (ICF) model, i.e., impairment (e.g. breath support, voice quality, loudness [23-28]), activity (intelligibility and naturalness, [24, 27, 29]) and participation [27, 28]. There is thus mounting evidence that speech intervention can have benefits both for speech and wider communication impact in people with progressive ataxias. However, most of the interventions have required relatively intensive input from clinicians, usually provided in individual patient settings. This is labour intensive and costly on the part of the health provider and can increase wait times for other patients.

An alternative model of care that addresses the demands on clinician’s time is group therapy. A recent systematic review on the benefits of this model in acquired dysarthria [30] found that it may increase treatment intensity and be potentially more cost-effective. The authors also highlight the increased opportunities for socialization, support, and integration of more client-driven goals into the activities, and how the practice in more naturalistic contexts as well as the social aspects of group intervention can facilitate better generalization of treatment targets and potentially also motor learning. More specifically, researchers have found that well-structured group therapy can provide similar quantitative benefits to individual therapy in primary intervention for speech deficits, as reflected by significant improvements in measures such as vocal intensity [31-35], maximum phonation time (MPT) [32, 33, 35, 36] and intelligibility [37, 38]. In addition, studies have demonstrated that group therapy can be effective in maintaining the gains resulting from intensive individual therapy [33, 39]. One aspect that has been highlighted as unique to group therapy is the social support patients provide to each other. This is reported to improve confidence and self-esteem [40-43]. In addition, participants may feel like they can contribute and participate more and tackle speech goals relevant to them in a more naturalistic settings [40, 44]. Such outcomes are particularly important in addressing the psychosocial impact reported in speakers with dysarthria such as loneliness and social isolation [17, 18, 20], which has been particularly exacerbated by the Covid-19 pandemic and the associated lockdown measures.

Whilst a number of studies suggest group intervention can lead to similar speech outcomes as individual therapy, this might come at a cost as studies have shown that higher dosage achieves better outcomes [45, 46], thus reducing the cost comparison between the two care models. To address this issue, we developed a novel treatment model - ClearSpeechTogether - that maximises treatment intensity whilst minimising clinician time. ClearSpeechTogether is a mixed individual— group therapy design. Its novelty lies in the fact that group sessions’ are facilitated by the patients themselves rather than trained clinicians, thus reducing pressure on health services whilst maximizing opportunities to internalize speech strategies for patients in a supportive, naturalistic environment.

This study aimed to establish the basis for future larger investigations by piloting the effects of ClearSpeechTogether on people with progressive ataxia and communication difficulties, and to establish the feasibility and acceptability of the approach. Our research questions were as follows:

RQ1: What is the feasibility of ClearSpeechTogether in a population of people with progressive ataxia?

Outcome measures: recruitment, attrition, adherence, need for additional individual sessions

RQ2: What is the acceptability of the approach to participants?

Outcome measures: fatigue measures, qualitative participant feedback on delivery format

RQ3: What are the potential communication and psychosocial benefits, and adverse effects of the approach?

Outcome measures: maximum phonation time, voice quality, intelligibility, sentence production consistency, communication participation, communication confidence, qualitative participant feedback, clinician observations

The study is reported according to CONSORT 2010 statement: extension to randomised pilot and feasibility trials [47].

## Materials and Methods

### Trial design

This 12-month study was a rater-blinded, single cohort design of patients with dysarthria due to progressive ataxia, using a single study arm – ClearSpeechTogether. Participants acted as their own controls by implementing a two week no treatment phase. No adjustments were made to the methodology following registration of this study in the ISRCTN clinical trial database [48].

### Sample Size

The study was intended to function as a pilot study to establish suitability of the intervention approach for a larger RCT. For this purpose, it was decided to run two groups of five participants each, aiming for a total of ten recruits for the study.

### Participants

Eligibility criteria for the study included a confirmed diagnosis of progressive ataxia, the presence of mild to moderate predominantly ataxic dysarthria, the absence of a functional voice disorder other than can be expected as part of the ataxia, the absence of visual, hearing or cognitive impairments that would have impact on participation in the assessment or treatment regime, age above 16 years, and availability and ability to use the technology necessary to complete assessment and treatment sessions online via Zoom.

Advertising took place via the funder website and social media campaigns. In addition, people with ataxia who had requested to stay informed about upcoming trials from a previous study were contacted directly via email. All participants self-selected and were provided with study information by email after contacting the research team for more information about the study. For those still interested, suitability to participate was established during a zoom call with the first author, during which consent was also taken for those recruited to the study.

### Study Design

Patient involvement in this study lasted for 16 weeks. This included a two-week pre-therapy assessment period, six weeks of intervention, and a further eight-week assessment period. Given the distance of study participants’ homes to the investigators and to each other, and the fact that the UK was undergoing various COVID-19 related lockdown measures at the time of the study, all assessments, individual and group therapy sessions were delivered remotely via Zoom. The feasibility of telehealth provision in this population had been established in our previous study using Skype [25].

Assessments required participants to record themselves at home. For this purpose, they were provided with information on how to use freely available recording software Audacity^R^ (version 3.0.3). Two participants had iPads and used the inbuilt voice recorder instead. Each participant was provided with a headset microphone to ensure stable mouth-to-microphone distance and a low-cost speech intensity meter (Cadrim Digital Sound Level Meter). They were sent a OneDrive link to securely upload their recordings after each assessment session. Backup recordings were made using Zoom cloud recordings following the participants’ consent.

### Assessment Tasks

The study included multiple baseline assessments (sessions 1 & 2, administered two weeks apart prior to treatment), and two post-therapy assessments, one within one week of completing treatment, and another 8 weeks post-treatment (Sessions 3 & 4). Assessments were conducted by the first author who was not involved in the treatment of participants.

In line with the ICF model, we assessed participant’s communication at impairment, activity and participation level. In addition, we collected information on fatigue and their medical history, as summarised in Table 1.

**Table 1:** Assessment Summary

| **Task** | **Session** | **ICF level** | **Measure** |
| --- | --- | --- | --- |
| Demographic and medical information | 1 |  | NA |
| Fatigue Impact Scale | 1 & 3 |  | Total score |
| Maximum phonation time | 1-4 | Impairment | Maximum duration in sec<br>Perceptual voice quality |
| Voice Quality | 1-4 | Impairment | GRBAS score |
| Reading passage | 1-4 | Activity | Intelligibility (DME) |
| Monologue | 1-4 | Activity | Intelligibility (percentage scale) |
| Sentence repetition | 1-4 | Activity | Utterance to utterance variability (UUV) |
| Communication Participation Item Bank | 1 & 3 | Participation | Total score (10 items, 0-3 scale) |
| Communication Confidence | 1 & 3 | Participation | Total score (1-10 scale) |
| Interview | 1 & 3 | Participation | Content Analysis |

Fatigue was captured with the overall score of the Fatigue Impact Scale (FIS [49]). For speech, two repetitions of maximum phonation time were collected unless the participant clearly performed within the normal range with a duration of around 20 seconds or more on their first attempt. Where two attempts were collected, the better of the two was used for subsequent analysis. Connected speech samples were captured by a reading task and a spoken monologue. The reading passage comprised the first paragraph of the Caterpillar passage [50], which resulted in 20 to 30 second samples, and for the monologue participants were asked to talk about a topic of their choice, such as a holiday, a recent memorable event or a hobby for about one minute. In addition, we recorded ten repetitions of the sentence “Tony knew you were lying in bed” to measure the consistency of sentence production (utterance to utterance variability, UUV [51]). This measure had previously shown some promise of being sensitive to intelligibility levels and could thus potentially quantify any post-treatment improvements in this parameter.

During the post-treatment assessment sessions, all participants were explicitly requested to apply the speech strategies developed during the intervention phase.

Finally, we captured participation by asking participants to complete the Communication Participation Item Bank (CPIB [52]) and to score their level of communication confidence on a 10 point scale.

### Analysis

The primary speech outcomes measures were duration and voice quality in the MPT, and intelligibility of connected speech. In addition, secondary outcomes, i.e., sentence production consistency, measures of participation, fatigue ratings and patient perceptions are also reported. All examiners were blinded to the time-point of the samples they analysed.

#### Vowel Prolongation

Vowel prolongation was analysed in terms of MPT and voice quality. Oscillographic and wide-band spectrogram data viewed in Praat ([44], version 6.0.43) were used for duration measures. In addition, four experienced SLTs used the GRBAS [53] to provide perceptual evaluations of voice quality. This tool provides scores for Grade (G - overall severity), roughness (R), breathiness (B), asthenia (A – weak voice) and strain (S).

#### Connected Speech

Intelligibility in the reading and monologue tasks were rated by four experienced SLTs. Due to the repetitive nature of the reading material, listeners scored the samples using the Direct Magnitude Estimation (DME) method [54]. This method uses a standard, which is given a score of 100, and asks listeners to rate a given speech sample in relation to this standard, where a score of 50 represents a sample half as intelligible or natural, and a score of 200 twice as intelligible or natural as the standard. We used the first recording of the reading sample (session 1) as the standard, listeners then assigned scores to the remaining three samples in comparison with the standard. These three samples were presented in randomised order. Listeners were blind to the status of any of the recordings. They were instructed to listen to the whole sample before scoring to account for potential variations in speech quality over time. The geometric mean was then calculated to arrive at an overall score per sample.

For the evaluation of the monologue, listeners scored the samples on a percentage scale. The samples comprised of around 30 seconds continuous speech without interruptions from the examiner or extraneous noise. As for reading, all samples were presented in randomised order of assessment but grouped by speaker.

#### Communication participation

We conducted semi-structured interviews in sessions 1, 3 and 4 to establish the form, severity and impact of speech problems experienced by participants, and how these were affected by the intervention. We also asked them to complete the short form of the Communication Participation Item Bank (CPIB) [52] on these occasions, and to provide a single score on a scale of 1-10 of their confidence when communicating with people outside their immediate social circle. Any changes in scores between sessions were discussed with participants to establish they truly reflected their perceptions.

#### Acceptability of the approach

The participant interviews also focused on the content and presentation of the treatment, discussing areas such as appropriateness of the exercises, treatment intensity, group dynamics, online nature of presentation and balance of individual versus group input.

### Inter-rater Reliability and Statistical Analysis

To assess inter-rater reliability, we conducted an independent analysis of four participants for the various measures performed. Agreement was excellent with an intraclass correlation coefficient of .999 for the MPT task. In addition, agreement between the four expert listeners for the perceptual analysis of the data was good with an ICC of .804 for reading intelligibility, and .884 for the monologue intelligibility, and .808 for voice quality.

Due to the small sample size and variability of speaker presentation non-parametric statistics were applied to avoid overinterpretation of results. The Friedman Test was performed to assess changes across time, using the Wilcoxon Signed rank test for the post-hoc analyses. We chose not to employ Bonferroni corrections given the sample size and highly exploratory nature of the investigation, but considered these factors in the interpretation of the results. For correlational analyses we employed Spearman’s Rho. Listener agreement was calculated with the Intraclass correlation coefficient.

### Treatment Schedule

Treatment was administered over a period of six weeks. This included an initial two weeks of individual therapy with two sessions of 45-60 minute per week (4 individual sessions) and twice daily homework tasks. This was followed by four weeks of intensive peer supported group practice, consisting of daily 1hour virtual meetings with the group (20 group sessions). The group phase was supported by a weekly meeting with the SLT. There was the option to provide further individual input for participants if the clinician determined that they were not using the speech strategies effectively or showed adverse reactions. A non-specialist volunteer was present during the non SLT-led group sessions to support the participants with any technical issues. The SLT sessions were administered by two expert clinicians who were highly experienced in treating patients with ataxia.

### Treatment focus

In line with previous trials for people with progressive ataxia [25, 27, 29, 55], two global speech strategies were focused on in this study – LOUD and CLEAR. Principles of the Lee Silverman Voice Treatment (LSVT) programme were adopted in terms of the focus on voice, however, unlike in Parkinson’s Disease, volume is not necessarily in issue for people with ataxia. Even though we maintained the cue “LOUD” for participants, it represented effective voice use and clinicians also ensured that voice quality was not strained or effortful, and produced at the appropriate pitch. “CLEAR” speech production aimed to maximize intelligibility for a communication partner by encouraging participants to over-articulate. The individual sessions were used to introduce participants to the two therapy concepts and to establish their use at least at single word level. The group phase then involved participants working through a handbook of graded exercises in line with the LSVT programme. These briefly involved further practice at the single word level and then moved quickly on to phrases, sentences and increasingly complex reading and free speech exercises by the end of week 4. Participants also practised ten daily phrases during the sessions, and completed prolonged vowel exercises as a warm up before the group meeting, to maximise time during the session for targeted speech activities.

Participants were provided with materials to practise, but also increasingly asked to prepare their own materials to build independence during the post-therapy phase for continued practice. The SLT met with the group at the end of each week to monitor each participant’s progress, suggest adjustments as necessary and to explain the upcoming tasks for the following week. The weekend was available for participants to prepare materials as necessary.

Participants were invited to reflect and comment on each other’s performance in a positive way. This was intended to provide support but also developed participants’ monitoring skills of themselves and others. All exercises were designed to be executed in turns, ensuring active involvement of all participants throughout the session. In addition, participants took turns in “chairing” each session, which meant they kept an eye on the time to ensure all exercises were attempted. They were also responsible for contacting the research team in case they were unclear about any of the exercises or any other problems arose.

Sessions tended to last 45 to 60 minutes depending on how much social chat was included at the start and end of the meeting. Speech exercises were designed to last 20 to 30 minutes.

## Results

### Recruitment

The recruitment period lasted 3 weeks. We had an unprecedented level of interest in the study, with more than 50 people with ataxia getting in touch either with the research team directly or Ataxia UK who had advertised the study through their various channels. Not all people provided details about themselves in the introductory email, and it is therefore difficult to ascertain how many of them would have been eligible for the study. Of those who provided relevant inclusion details, a small number were not eligible, either due to other comorbidities such as myasthenia gravis, or due to their geographical location, such as residency in the US or Canada. The remaining candidates were contacted in the order they approached us until all ten places were filled. One person had to be rejected due to a co-morbidity presenting during this process.

### Baseline Data

Twelve patients with progressive ataxia were recruited. Their details are summarised in Table 2, including medical history and dysarthria features. As can be seen, ataxia diagnosis varied considerably across participants. Due to the movement restrictions and long waiting lists to see health professionals imposed by COVID19 lockdown no up to date neurological examination was conducted as part of this study. Instead, we relied on patient reports of their medical history and used a broad grading of their motor disability as stage 0 = no gait difficulties, stage 1 = disease onset, as defined by onset of gait difficulties; stage 2 = loss of independent gait; stage 3 = confinement to wheelchair; stage 4 = death [56] which was deemed sufficient for the purposes of this pilot study which focused entirely on their speech performance. The severity of the latter was derived from the intelligibility ratings of the monologues. The majority of our participants were rated as showing mild to moderate gross motor impairment (stages 1 & 2, Table 2). In addition, most had a mild to mild-moderate level of speech impairment, with only two located at the lower moderate to severe end of the spectrum (AD8 & AD9, Table 2).

**Table 2:** Participant Characteristics

| Participant | Age range | Gender | Diagnosis | Years since diagnosis | Motor impairment | Intelligibility deficit (% scale) |
| --- | --- | --- | --- | --- | --- | --- |
| AD1 | 56-60 | M | SCA28 | 2 | 1 | 74 |
| AD2 | 65-70 | M | Idiopathic<br>Cerebellar<br>Ataxia | 6.5 | 1 | 71 |
| AD3 | NA | NA | NA | NA | NA | NA |
| AD4 | 56-60 | M | Idiopathic<br>cerebellar ataxia | 1 | 2 | 78 |
| AD5 | 60-65 | F | CANVAS | 9 | 2 | 75 |
| AD6 | 66-70 | M | Idiopathic<br>cerebellar ataxia | 1 | 1 | 79 |
| AD7 | NA | NA | NA | NA | NA | NA |
| AD8 | 46-50 | M | SCA3 | 17 | 3 | 45 |
| AD9 | 56-60 | M | Presumed<br>Autoimmune<br>ataxia | 3 | 2 | 51 |
| AD10 | 56-60 | F | Friedreich's<br>Ataxia | 10 | 2 | 66 |
| AD11 | 66-70 | F | SCA6 | 5 | 2 | 69 |
| AD12 | NA | NA | NA | NA | NA | NA |

### Adherence

Of the twelve patients recruited, eleven commenced and nine completed treatment (Table 3). Retention was not impacted by the intervention approach, but rather significant personal circumstances in both cases who had to discontinue and both expressed keen interest in re-joining at a later date. In the first case (AD3) we were able to fill the place with AD6. AD7 took part in the 2-week individual therapy phase, but had to drop out once the group sessions began. In order to maintain the same group size, we admitted a further participant, AD12, to join the group sessions in place of AD7 on day 2 of the group phase. This was possible as he had very mild dysarthria and was able to internalise the speech strategies very quickly after only one individual session. He was closely monitored by the SLT in case further sessions were needed. As he had not received the four individual sessions or the two assessment sessions, his data are not reported in this study. Adherence to treatment was generally good. Three participants (AD5, AD8 and AD11) experienced several days gaps in participation in the first week of the group phase due to illness or personal circumstances. They attended all remaining sessions.

**Table 3:**
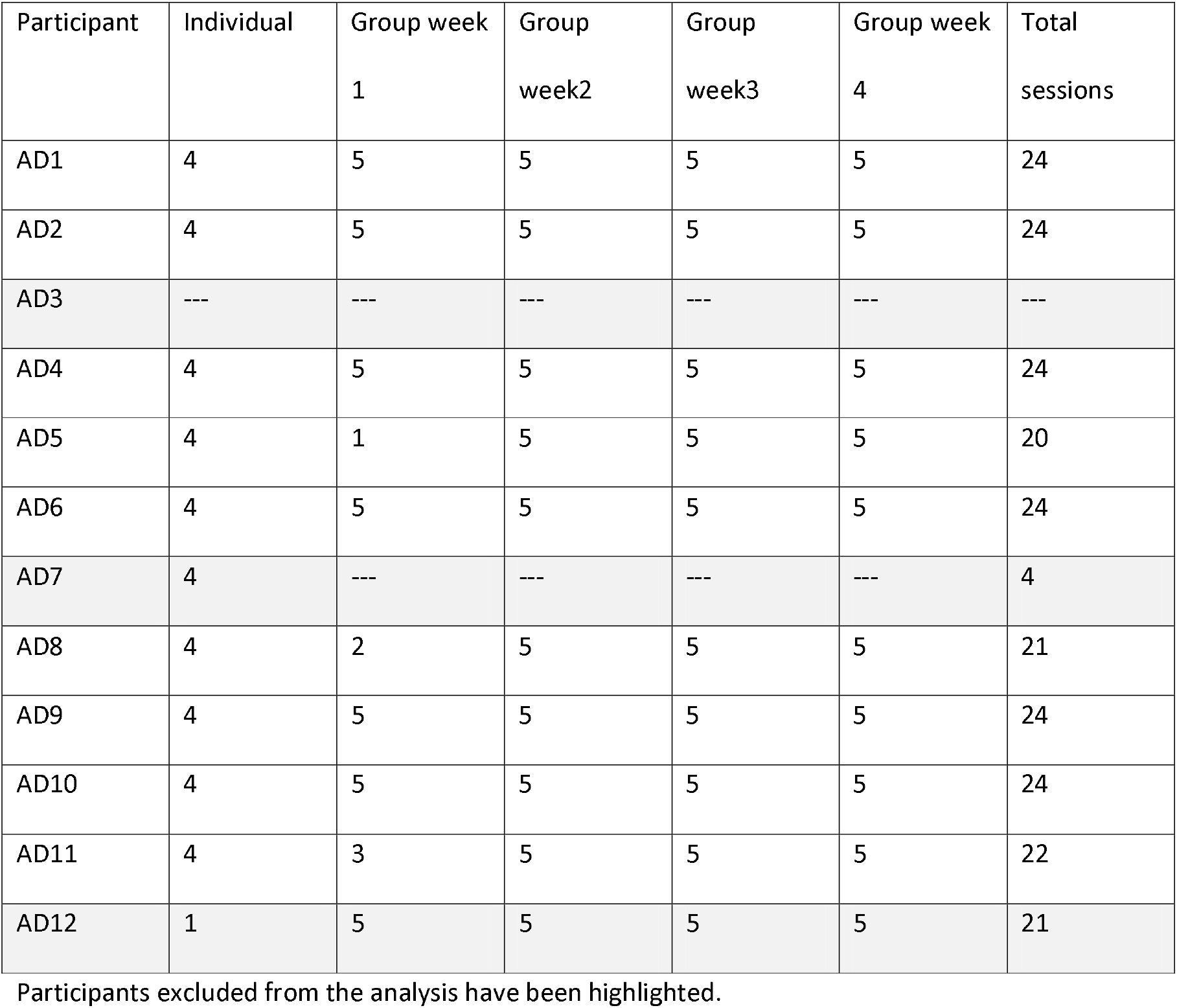
Treatment Adherence (expected attendance at 4 individual sessions and 5 weekly group sessions)

| Participant | Individual | Group week<br>1 | Group<br>week2 | Group<br>week3 | Group week<br>4 | Total<br>sessions |
| --- | --- | --- | --- | --- | --- | --- |
| AD1 | 4 | 5 | 5 | 5 | 5 | 24 |
| AD2 | 4 | 5 | 5 | 5 | 5 | 24 |
| AD3 | --- | --- | --- | --- | --- | --- |
| AD4 | 4 | 5 | 5 | 5 | 5 | 24 |
| AD5 | 4 | 1 | 5 | 5 | 5 | 20 |
| AD6 | 4 | 5 | 5 | 5 | 5 | 24 |
| AD7 | 4 | --- | --- | --- | --- | 4 |
| AD8 | 4 | 2 | 5 | 5 | 5 | 21 |
| AD9 | 4 | 5 | 5 | 5 | 5 | 24 |
| AD10 | 4 | 5 | 5 | 5 | 5 | 24 |
| AD11 | 4 | 3 | 5 | 5 | 5 | 22 |
| AD12 | 1 | 5 | 5 | 5 | 5 | 21 |

Participants excluded from the analysis have been highlighted.

### Numbers Analysed

Nine participants completed the full programme, and eight of these were included in the analysis. AD11 was diagnosed with fluid in her lungs after the individual therapy had been completed. Her speech data are therefore not considered in the group statistics as her pre-treatment performance would have been impacted by the presence of the fluid. In addition, we experienced problems with data quality in the final assessment of AD8 which affected his own and the backup recordings. Consequently, none of his assessment 4 data were considered with the exception of his MPT performance which could be reliably extracted.

### Outcomes

#### Fatigue

Global fatigue scores from the FIS ranged from 4 to 10 (with ten being normal) at first assessment, with a mean of 6.3 (Table 4). Comparison with scores at assessment 3 suggested no significant change (*p* = .680). Where scores did change (three participants), this was to the better.

**Table 4:**
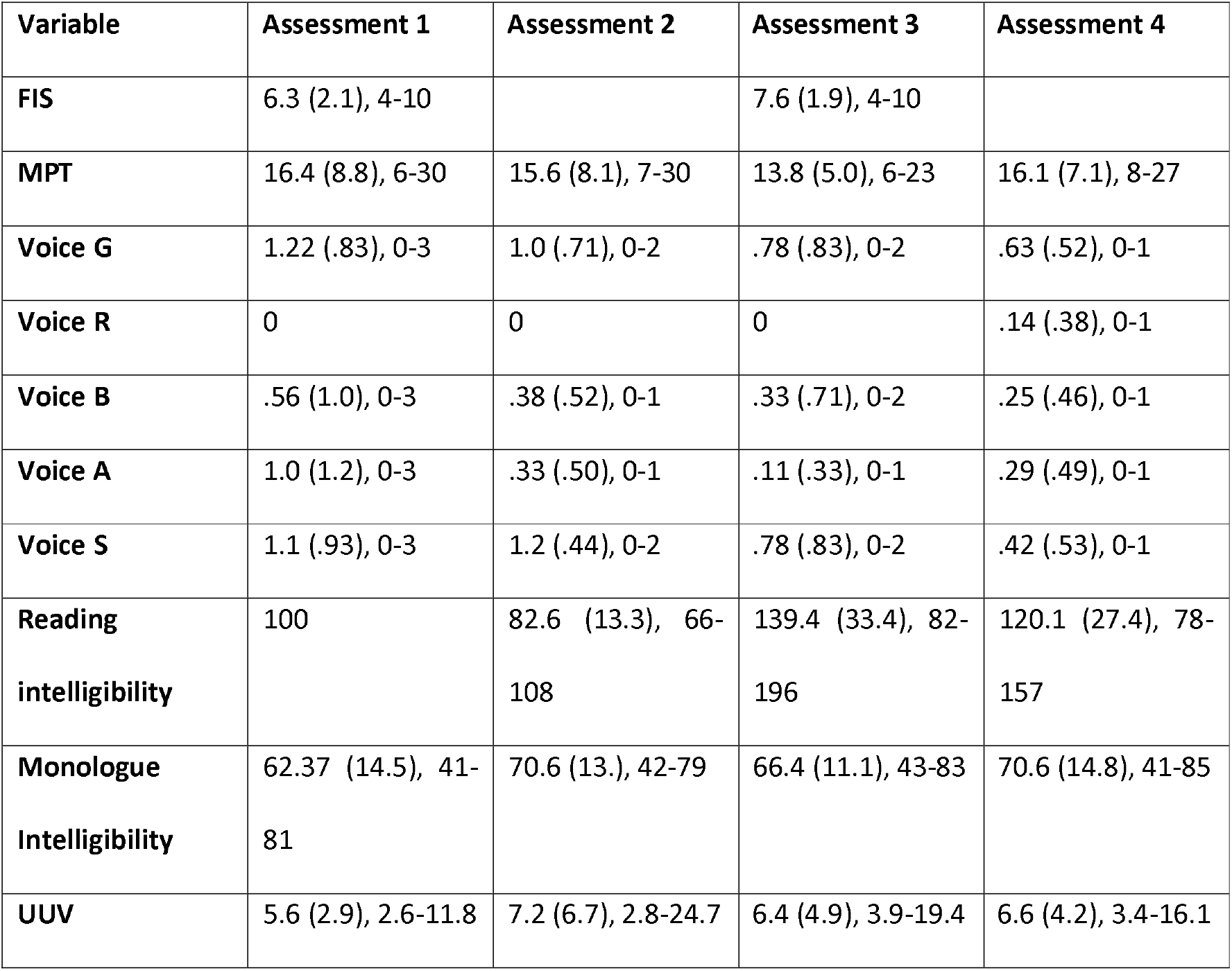
Means, Standard Deviations and Range of Speech Variables

| Variable | Assessment 1 | Assessment 2 | Assessment 3 | Assessment 4 |
| --- | --- | --- | --- | --- |
| FIS | 6.3 (2.1), 4-10 |  | 7.6 (1.9), 4-10 |  |
| MPT | 16.4 (8.8), 6-30 | 15.6 (8.1), 7-30 | 13.8 (5.0), 6-23 | 16.1 (7.1), 8-27 |
| Voice G | 1.22 (.83), 0-3 | 1.0 (.71), 0-2 | .78 (.83), 0-2 | .63 (.52), 0-1 |
| Voice R | 0 | 0 | 0 | .14 (.38), 0-1 |
| Voice B | .56 (1.0), 0-3 | .38 (.52), 0-1 | .33 (.71), 0-2 | .25 (.46), 0-1 |
| Voice A | 1.0 (1.2), 0-3 | .33 (.50), 0-1 | .11 (.33), 0-1 | .29 (.49), 0-1 |
| Voice S | 1.1 (.93), 0-3 | 1.2 (.44), 0-2 | .78 (.83), 0-2 | .42 (.53), 0-1 |
| Reading intelligibility | 100 | 82.6 (13.3), 66-108 | 139.4 (33.4), 82-196 | 120.1 (27.4), 78-157 |
| Monologue Intelligibility | 62.37 (14.5), 41-81 | 70.6 (13.), 42-79 | 66.4 (11.1), 43-83 | 70.6 (14.8), 41-85 |
| UUV | 5.6 (2.9), 2.6-11.8 | 7.2 (6.7), 2.8-24.7 | 6.4 (4.9), 3.9-19.4 | 6.6 (4.2), 3.4-16.1 |

#### Maximum Phonation Time

Maximum phonation time at first assessment was variable across participants (Table 4) and no post-treatment effects could be identified (*p* = .366, χ2= 3.17, df=3). Normative data on MPT varies considerably in the literature (see reviews by [57, 58]) and is influenced by both the age and gender of the participants. For the age range of the current participants, group means tend to sit around 15 seconds. Using this cut off, only two participants fell below the normal range in the current group, and both showed a small increase in performance, moving from 6 to 13 sec (AD5) and 9 to 13 sec (AD10) from assessment 1 to assessment 4. The remaining speakers achieved MPTs of up to 30 seconds pre-treatment, and no further improvement was therefore expected.

#### Voice Quality

Most dimension on the GRBAS showed no significant changes across the assessment points. However, strain was significantly reduced between pre-treatment and the 8 week follow up (Friedman test: *p* = .010, χ2 = 11.43, df = 3; post-hoc tests: A1-2: p= 1.000, A1-A3: p = .257, A1-A4: p = .180, A2-A3: p = .034, A2-A4: p = .014, A3-A4: p = .317).

#### Reading & Monologue Intelligibility

The Friedman tests for the reading data suggests significant changes across the four assessment sessions (Friedman test: *p* = .001, χ2 = 16.54, df=3). post-hoc tests: A1-2: p= 1.000, A1-A3: p = .257, A1-A4: p = .180, A2-A3: p = .034, A2-A4: p = .014, A3-A4: p = .317). Post-hoc comparison between the two pre-treatment sessions showed that intelligibility ratings were significantly lower in the second assessment (p = 0.28). On the other hand, intelligibility was significantly increased from both pre- to immediately post-treatment (A1-A3: p = .018, A1-A4: p = .018, A2-A3: p = .128, A2-A4: p = .018). Performance decreased again to some degree from A3 to A4, although not significantly (p = .091). The decrease in intelligibility resulted in comparisons between A1 and A4 to be non-significant, however, the intelligibility level remained higher in A4 than pre-treatment for most participants with the exception of AD1 and AD10 (Figure 1). AD8 was the only speaker to not show improvement after therapy.

**Fig. 1.**
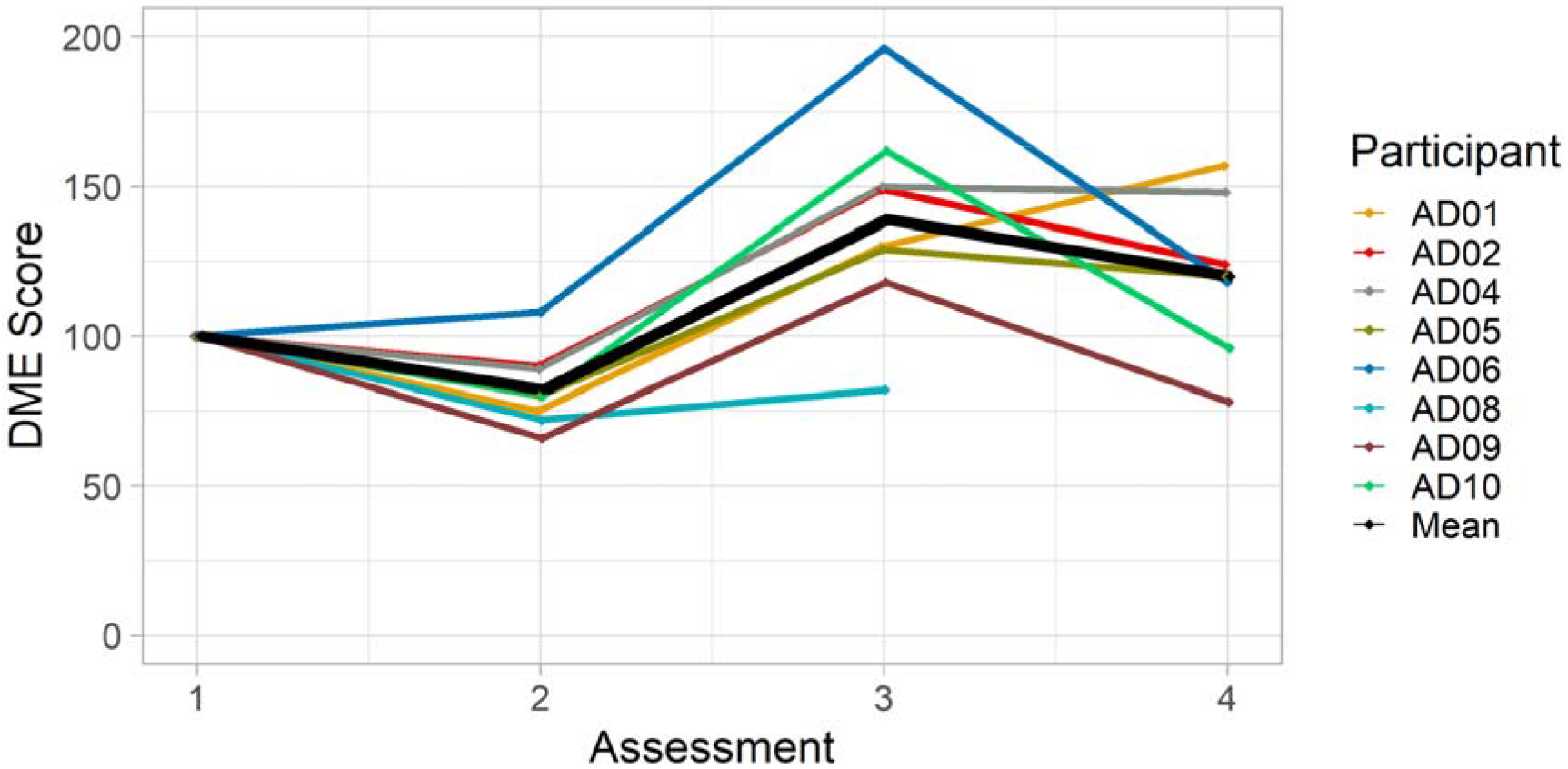
Reading Intelligibility across assessment session by speaker and group mean

In contrast to the reading data, the monologue did not show changes between any of the assessment points (Friedman test: *p* = .125, χ2 = 5.75, df=3). Descriptive analysis indicated that three of the nine participants had some higher scores in at least one of the post-treatment assessments, but this was not sufficient to return a statistically significant result.

#### Utterance to utterance variability

There was no significant difference in UUV values across the assessments (Friedman test: *p* = .968, χ2 = .257, df=3). Performance was highly variable with no trends identified. There was also no correlation between the UUV values and reading intelligibility where treatment effects had been present (Spearman’s rho for cumulative data for Assessment 2 – Assessment 4: *r* = .027, *p* = .882). The UUV measure does therefore not appear suitable to track progress after treatment.

#### Communication participation and confidence

Participants completed the CPIB [52] and their rating of communication confidence once pre-therapy and at each of the post-treatment assessment points. The data for the CPIB (Figure 2) show that each participant rated the impact of their dysarthria on communication participation as being reduced immediately post-treatment. In four cases, participation continued to improve up to 8 weeks after treatment. The same number of participants maintained the reduced levels post therapy. Only one person (AD8) reported a worsening of impact longer term, although it was still perceived as lower than pre-therapy. Discussion with him revealed that he had been staying with his extended family at the time of assessment 3, but then returned home where he lived in relative isolation during the period leading up to assessment 4. This had reduced his ability to participate in communication, hence the lower scores. As expected, the statistical analysis returned significant differences between pre- and post-treatment values and no difference between immediate and long-term post-treatment assessments (Friedman test: *p* < .001, χ2 = 15.70, df=2; post-hoc tests: A1-A3: p = .007, A1-A4: p = .008, A3-A4: p = .141).

**Fig 2.**
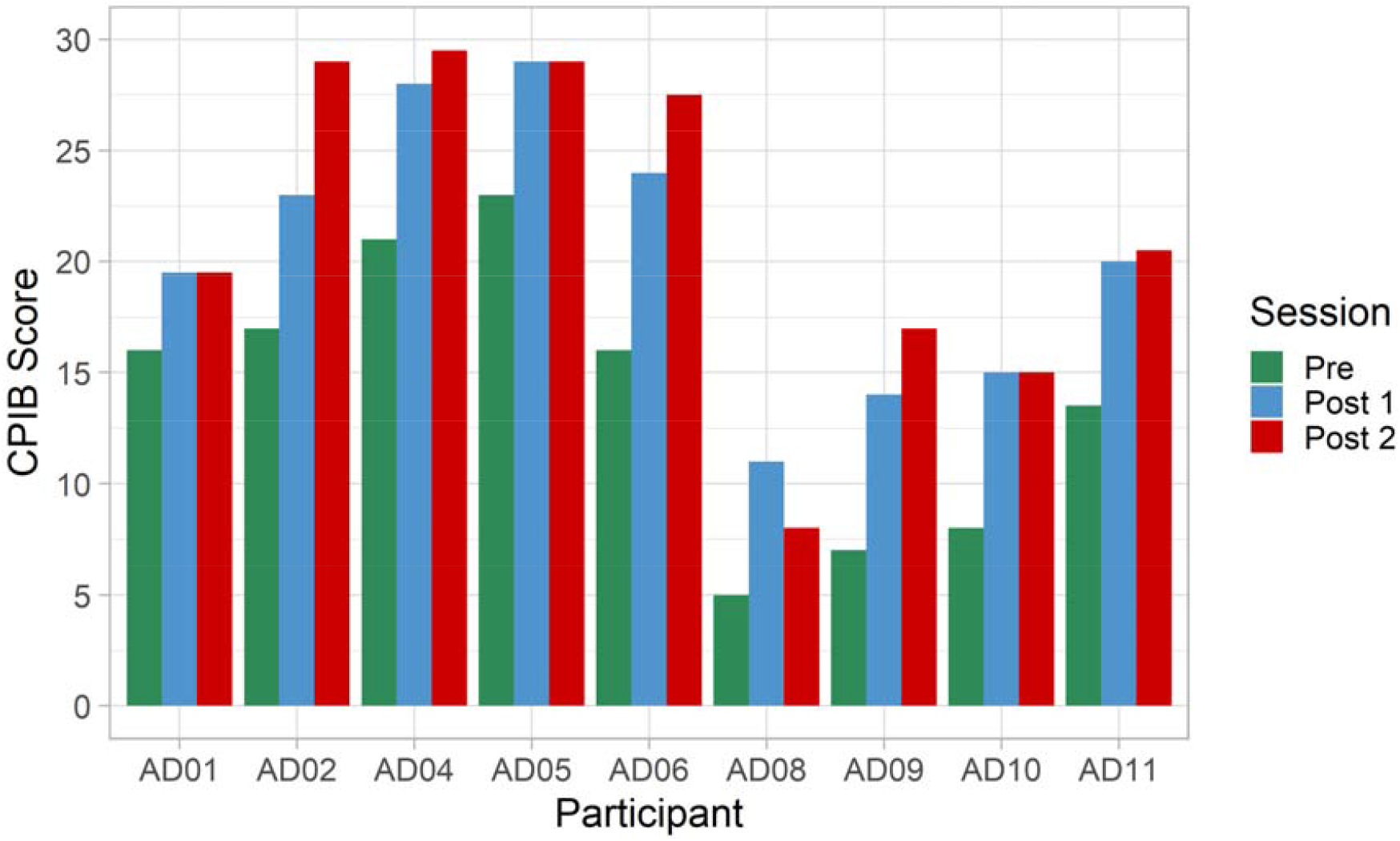
Communication Participation Item Bank (CPIB), maximum scores (no impact) = 30

A similar picture is presented by the confidence ratings (Figure 3), i.e., all but one participant increased their confidence post-treatment, and most maintained or further increased their rating 8 weeks later. The participant who showed no change (AD5) had rated her confidence at the highest level pre-therapy, therefore no further improvement could be expected. Two participants, AD8 and AD10, reported reductions 8 weeks post-treatment, both attributed the change to less opportunities to communicate with people outside their immediate family and friends circle due to renewed COVID 19 lockdown restrictions. The other participants varied in the degree of change post-treatment, with some reporting considerable increases (e.g., AD2 from 3.5 to 7.5, or AD9 from 1.5 to 5). AD8’s positive change is noteworthy as he was the only speaker who showed no improvement in the intelligibility measures after treatment, suggesting that therapy could have positive impact in the psychosocial domain in the absence of changes to speech performance. Statistical tests confirmed the above observations (Friedman test: *p* < .001, χ2 = 13.15, df=2; post-hoc tests: A1-A3: p = .011, A1-A4: p = .018, A3-A4: p = .414).

**Fig. 3.**
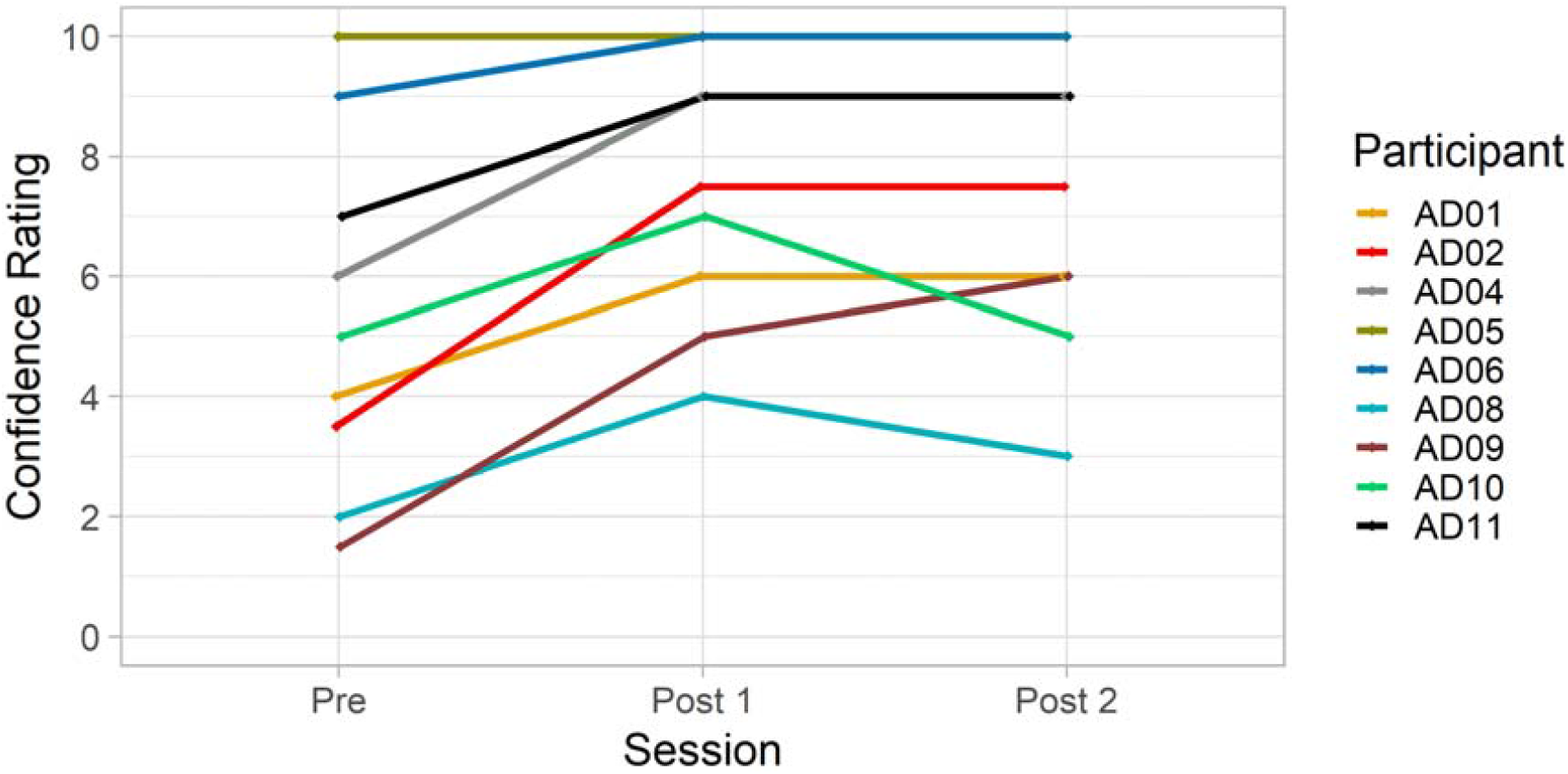
Confidence ratings pre- and post-therapy by participant (1= no confidence, 10 − full confidence)

Statistical analysis indicated that communication participation and confidence were correlated (assessment 1: r = 704, p = .034; assessment 3: r = .867, p = .002; assessment 4: r = .886, p = .001).

The relationship between these two aspects and speech severity as expressed by monologue intelligibility was more complex and changed over time. Participation was significantly correlated with intelligibility pre-therapy (r = .886, p = .019), as well as in assessment 4 (r = .780, p = .039), but not in assessment 3 (r = .716, p = .070). The relationship between confidence and intelligibility showed the reverse with no correlation at baseline (r=.589, p = .219) or assessment 4 (r = .588, p = .165), but a significant result at assessment 3 (.864, p = 0.12). Given the small participant numbers included in this analysis it appears that for most participants there was some relationship between the three aspects, but individual differences might have rendered results non-significant at any given assessment.

#### Participant Self-Perception of their Communication

When participants were asked in assessment 3 how their communication had changed, all reported positive outcomes of the intervention. Similar to our quantitative outcome measures, questions focused directly on their speech behaviour as well as their overall communication impact and confidence. A number of themes developed from the interviews:

Several participants mentioned that the voice had become “stronger” and that they were generally speaking louder than before. In addition, comments referred to the fact that voice had become more stable without sudden bursts in loudness. One participant had complained of flat, monotonous intonation before which she felt had been resolved by the intervention – “I don’t sound like a robot anymore” (AD11).

Participants appeared to have integrated the concept of clear speech well. They talked about speaking more clearly and deliberately after therapy, over-articulating, or trying to pronounce every syllable.

The clear speech strategy also impacted on pacing of speech and pausing, participants talked about speaking slower, spacing words out, and taking more breaths.

Effort was mentioned in a variety of different contexts. The effort required to speak was reported to be lower after therapy, allowing participants to speak in longer utterances and for longer periods of time – “I’m now quite happily chatting away to the hairdresser for 30 minutes” (AD6). They also reported improved ways of managing the effort they put into speaking, and an increased motivation to rally themselves despite being tired, which allowed them to participate more, in particular at times when they would normally have withdrawn from communication.

Similar to the results of the CPIB and confidence ratings, participants reported that they had increased their levels of communication confidence in general, or in particular speaking situations such as in larger groups or on the phone.

#### Participant Feedback on the Therapy Process

The interviews also explored a number of issues regarding the intervention regime, including the balance between individual and group sessions, intensity of delivery, content/treatment focus, and views about the group element of the intervention.

Participants felt generally positive about the intervention regime, indicating that it had addressed their needs and that they had a good understanding of the purpose and benefits of the strategies conveyed. They felt the balance between individual and group elements was good, which corroborated the clinician’s indication that most participants were able to move beyond the single word stage onto short as well as longer phrases during the individual therapy phase. The two more severely affected participants indicated they would have preferred additional individual input before commencing the group sessions. This had in fact been picked up by the SLT and they were each offered a further session in week 2 of the group phase. The additional session resolved the issues from the participants’ as well as clinician’s view. Scheduling had not been a problem as no one was in employment and other activities were reduced due to the COVID 19 lockdown restrictions. Some indicated that the intensity of the group sessions might have been an issue if they had had other parallel commitments such as gym sessions, or had been in work. The online nature of the intervention was not seen as a barrier and welcomed by many as it facilitated participation. Group dynamics had been good in both groups and members continued to meet on their own accord twice a week after the intervention was complete for social reasons and to continue to practice together. Participants also listed a range of benefits related specifically to the group meetings which could be categorised as social and speech benefits, as outlined in Table 6. Overall, participants were highly positive about the group activity. Whilst the social benefits described could also have been achieved by attending a charity support group, the speech benefit were specifically related to our therapy model. Two participants who had recently attended individual SLT indicated that the group meetings had added benefits for them. In addition, a further two participants commented that having to lead the groups themselves helped reinstate former social roles, e.g., they felt more in charge again.

**Table 6:** Themes related to group intervention benefits

| **Social Benefits** | **Speech Benefits** |
| --- | --- |
| Meeting other people with ataxia | Opportunity to talk, everybody gets a turn |
| Feeling you're not the only one who has the problem | Feedback from others / constructive criticism valuable to learning |
| Find out more about different ataxia presentations and severities and associated problems | Taking cues from one another / hearing others use speech strategies helps and motivates to integrate them into own speech |
| Sharing coping strategies | Helps conquer apprehension about speaking in a more supportive environment / gives confidence talking with others |
| Sharing frustrations | Improves motivation to practise |
| Psychological support | Allows practice of real-life speaking |
|  | situations, re-establishment of roles |

## Discussion

The aim of this study was to pilot the feasibility, acceptability and potential effectiveness of ClearSpeechTogether in a small group of people with progressive ataxia and mild to moderate dysarthria.

### Feasibility

Recruitment was highly successful with a large waiting list of participants remaining. Retention to the study was 80% with reasons for dropping out of the study based on significant personal circumstances. Adherence was also good. Given the length of the intervention period it was inevitable for some participants to miss some of the group sessions due to illness or other reasons. Two thirds of participants attended all sessions, the remaining third missed 20% of group sessions or less. These figures compare favourably with our previous study [25] which experienced issues of recruitment and adherence. One major difference in the current study was that recruitment was not restricted to people with FRDA. Although this group is highly likely to experience communication problems, a recent survey suggests that these may have a lower impact on people than for other types of progressive ataxias [21]. This could explain the lower interest in participating in our previous trial targeted at FRDA. Furthermore, the impact on communication opportunities as well as access to standard SLT provided by the health service caused by the COVID19 lockdowns could have motivated more people to volunteer for the study this time round.

### Acceptability

Qualitative data indicates that participants found the programme addressed their communication needs, that the content, scheduling and delivery of treatment were appropriate, and that the group element had added benefits for them. No adverse effects were reported from the speech treatment and fatigue did not change significantly. Group dynamics were also positive, to the degree that groups continued to meet socially online and later face to face after the completion of the study. Our study thus concurs with previous reports of psychosocial benefits of group intervention (e.g., [41, 42]).

From a clinician’s point of view, the current model translated into an average time commitment of 5-6 sessions per patient, comparable to standard NHS input. This compares favourably with the 24 sessions the patients participated in. Some additional help was provided by the non-clinical volunteers to ensure the group settled and experienced no technical difficulties during the Zoom calls. Although the volunteers remained in place for the entire group phase for this pilot investigation, they indicated that the participants would have managed by themselves after some time. The exact duration and nature of support required needs to be determined in larger trials. There were no access issues related to the online provision in our study, and all participants were able to handle the necessary technology without aid. This aspect will need to be monitored in future to ensure equity of access to those who require treatment.

### Effectiveness

In relation to communication outcome measures, the pilot results indicate little change at the physiological level, some improvements in intelligibility post-therapy across at least some tasks and participants, and consistent improvements at the psychosocial level in relation to communication participation and confidence.

The lack of improvement in relation to maximum phonation time stands in contrast with previous trials focusing on this aspect [25, 26, 59]. There are several possible explanations for this outcome. First, the current study devoted less time to phonatory practice than is featured in LSVT and focused more on clear speech. In addition, these exercises were treated as a warm up activity, potentially resulting in less focus and effort on this task. Finally, the participants’ baseline performance mostly fell within the normal range, thus allowing less room for improvement. Importantly, the treatment was successful in increasing MPT where a clear impairment was present. In relation to voice quality, our sample generally had no or very mild impairment. This is similar to previous reports in the literature [12, 25, 27, 28]. Interestingly, in direct contrast to our previous study where all dimensions except strain showed some improvement post-treatment [25], strain was the only aspect to improve significantly this time. This might be explained by a different baseline voice profile of the participants and the difference in aetiology.

The results for the intelligibility component were promising for reading and in line with other therapies [24, 27, 29]. The results were less clear for the monologue task where only some speakers showed improvements and group statistics were not significant. Nevertheless, the current study provides preliminary evidence that the programme was successful in improving speech performance in at least some of the participants, warranting further investigation. The two participants with the least improvement in intelligibility had the most severe level of dysarthria. Whilst they were able to work through the programme alongside the other participants, they required additional individual sessions as they did not apply the speech strategies as effectively during the exercises. Little can be said on the basis of only two participants, however, future studies should investigate whether more severely affected speakers require a higher proportion of individual therapy or a slower progress in task demand to allow them to fully internalise the strategies before moving to the next level of complexity.

Most encouraging were the results of the analysis of psychosocial dimension. All participants reported improvements in communication confidence as well as participation. It was positive to see some participants develop this further post-treatment. Only two participants reported a decline longer term, and in these cases this could be directly linked to external events, e.g., one participant having moved from a supportive family environment with ample opportunity to communicate to living in isolation. Similarly, the other participant also mentioned negative impact of renewed COVID19 lockdown measures on her communication confidence. These findings highlight the importance of day-to-day opportunities to interact with others on long term effectiveness of communication performance. Another important finding was that although intelligibility and participation ratings were broadly related in line with Borrie et al.’s [60] findings, this was not always the case. For example, participants who dropped in intelligibility in assessment 4 still maintained their participation benefits, and AD8, who showed no notable improvements in intelligibility, reported a considerable increase in confidence and participation. This shows that no reliable assumptions can be made on day-to-day communication behaviour purely from clinical examination of speech. The results also raise the question whether intelligibility remains the most appropriate outcome measure for patient management and clinical trials or whether this should be replaced with measures that better reflect everyday communication behaviour of participants or the impact their speech problem have on them.

Finally, the qualitative evaluations stressed the added benefits the group treatment provided for both speech and psychosocial factors, similar to previous research on group interventions (cf. Whillans et al. [30] for a review). In addition, we identified specific benefits of the peer led design as reflected in the reports of positive changes to social roles in terms of having to take charge of the session. Social role limitations were highlighted as the second most impactful problem in people with FRDA in a recent interview study [61]. These can be difficult to address directly in a one-one-therapeutic context, rendering our result particularly encouraging in this respect.

## Limitations

Whilst the above reported outcomes are generally positive, it has to be stressed that they are based on a limited number of study participants’ experiences. The treatment model therefore requires to be further evaluated in a larger scale and sufficiently powered trial. In addition to potential speech and psychosocial benefits, the practical and health economic aspects also need to be investigated further in terms of feasibility of setting of treatment groups that are sufficiently matched and available at appropriate times to run the proposed model. The fact that the therapy schedule focused on relatively generic speech strategies should make it appropriate for a wider range of patients beyond those with progressive ataxia. In addition, whilst the participants with greater severities were able to complete the exercises at the same rate as the other group members, they did not improve their intelligibility to the same degree and required additional sessions. Future studies thus need to consider the impact of severity as well as type of disease to assess the effectiveness of ClearSpeechTogether as a generic intervention for people with acquired dysarthria.

## Conclusion

In conclusion, this study provides further support for the benefits of SLT for speakers with progressive ataxia across all ICF dimensions, and confirms earlier reports on the (added) value of group intervention. In addition, our model is cost-effective, providing intensive delivery of 24 client sessions over 6 weeks on the basis of only five to six clinician sessions, thus maximising the input currently available within the confines of the NHS. Telehealth delivery minimises the impact of the scheduling intensity during the group phase.

The current study thus provides an encouraging basis for further research into speech treatment in speakers with progressive ataxia as well as other related speech disorders, and into new models of treatment delivery that reduce the workload pressures of clinicians whilst ensuring high quality, effective treatment of the required intensity for patients.

## Data Availability

All data produced in the present study are available upon reasonable request to the authors

## Acknowledgements

This study was funded by Ataxia UK, grant reference ZSTRATHC. We would like to thank all our participants for their valuable time. Many thanks also go to the Ataxia UK team, James Atkins and Anastasia Georgousis, who supported the group sessions. Finally, we are grateful to our expert SLT listeners, Rebecca Kimber, Caroline Waszkiewicz and Conor Brown for their time to evaluate the data.

## Author Contribution

AL designed and managed this study, conducted assessment, analysed part of the data and authored this article; JC was involved in recruitment, sessions scheduling and preparation, and data analysis; ML and JG provided the treatment for the study; AE was involved in the planning of the intervention; FB analysed some of the data; and MH provided guidance during recruitment and contributed to the article.

## Conflict of Interest

The authors have no conflicts of interest or financial or non-financial interests to disclose.

## Ethics Approval

This study was performed in line with the principles of the Declaration of Helsinki. Approval was granted by the Ethics Committee of the University of Strathclyde on 17.12.2020 (No. UEC20/92).

## Notes

### Competing Interest Statement

The authors have declared no competing interest.

### Clinical Trial

ISRCTN93368860

### Author Declarations

The ethics committee of Strathclyde University gave ethical approval for this work

